# Sputum alarmin levels delineate distinct T2 cytokine pathways and patient subgroups in asthma

**DOI:** 10.1101/2022.05.29.22275711

**Authors:** Samir Gautam, Jen-Hwa Chu, Avi J. Cohen, Ravdeep Kaur, Gabriella Wilson, Qing Liu, Jose Gomez, Haseena Rajaveen, Xiting Yan, Lauren Cohn, Brian J. Clark, Geoffrey Chupp

## Abstract

**Rationale:** Asthma is a chronic airway disease driven by multiple immunologic pathways that determine the clinical response to therapy. Current diagnostic methods are incapable of discriminating subtypes of asthma and guiding targeted treatment. We hypothesized that sputum cytokine profiles could help to identify immunologically-defined disease subtypes and individualize therapy in patients with severe asthma.

**Objectives:** Define asthma subtypes associated with sputum alarmin and cytokine levels.

**Methods:** Cross-sectional analysis of clinical features and sputum from 200 asthmatic patients was performed. 10 cytokines belonging to alarmin, T2, and non-T2 pathways were measured. Pearson correlation was used to identify cytokine modules. Latent class analysis was used to cluster patients by cytokine expression.

**Measurements and Main Results:** Three modules of highly correlated cytokines were identified including a non-T2 module, the IL-1β_mod_ (IL-1β, IL-6, GCSF), and two distinct T2 modules: TSLP_mod_ (TSLP, IL-4, IL-5, IL-9) and IL-33_mod_ (IL-33, IL-13, IL-21). The TSLP_mod_ was associated with asthma severity, airway obstruction, eosinophilia, and elevated FeNO. Patient clustering revealed three subgroups; two different subgroups showed expression of T2 modules.

**Conclusions:** Analysis of sputum cytokines revealed three discrete signaling modules in patients with asthma. Unexpectedly, the inclusion of alarmins led to separation of canonical T2 cytokines into two unique modules; IL-5 grouped with TSLP, while IL-13 grouped with IL-33. In addition, patient clustering revealed two distinct endotypes associated with T2 immune signaling. These findings indicate a new layer of immunologic heterogeneity within the T2 paradigm, and suggest that sputum cytokine profiling may hold diagnostic utility for patients with asthma.

## Introduction

Immune-mediated asthma was once considered a unidimensional allergic process characterized by T2 cytokines (e.g. IL-4, IL-5, IL-13), immunoglobulin E (IgE) production, eosinophilic infiltrates, atopy, airway hyperresponsiveness, and high FeNO. However, asthma is now recognized as a heterogeneous disease associated with diverse clinical characteristics (phenotypes) and pathobiologies (endotypes).(1) Patients demonstrate variable responses to medical therapy, likely due to differences in their immunologic endotypes.(2) A central question in the management of asthma is how to diagnose these endotypes and tailor therapeutic strategies accordingly.

The prevailing paradigm divides asthma into two categories: T2 and non-T2 disease.(3) The latter is associated with neutrophilic inflammation, fixed airway obstruction, smoking, obesity, and resistance to glucocorticoids.(4–6) Supervised analyses of patients in multicenter studies using clinical features (e.g. atopy) and commonly-used biomarkers (e.g. blood eosinophil levels) support the existence of T2 and non-T2 endotypes.(7–9) In practice, however, it is challenging to distinguish these endotypes and predict medication responsiveness based on standard clinical and laboratory metrics alone.

An alternative approach is to subclassify asthma according to molecular characteristics using ‘omics technology. Transcriptomic studies of sputum-derived cell samples, for example, effectively discriminate T2 vs. non-T2 disease, and have provided valuable insights into asthma pathophysiology.(10, 11) Metabolomic and proteomic analyses of airway samples have also been used to delineate patient subtypes.(12, 13) However, the technical demands and cost of these ‘omic approaches preclude their widespread clinical use.

In contrast, molecular endotyping of patients using sputum cytokines offers several potential advantages. First, sputum cytokine measurement is non-invasive, technically facile, and relatively low-cost. Second, since cytokines are central mediators of asthma pathogenesis, their characterization has the potential to yield insights into immunologic mechanisms of human disease.(13–16) Third, cytokines represent important therapeutic targets per se; thus, their levels could be used to determine personalized treatment strategies for patients. Biologic therapies targeting the T2 cytokines IL-4 and IL-5, for example, have been studied in depth over the last two decades.(17) More recently, trials have demonstrated the efficacy of inhibiting alarmins such as TSLP and IL-33 – master upstream regulators of T2 cytokine expression.(18–21)

In this study, we sought to characterize the expression of cytokines in the sputum of patients with asthma. We hypothesized that cytokine profiling may offer a novel, technically feasible means to subtype patients and inform the selection of anti-cytokine biologic therapies. In contrast to prior proteomic studies,(13) we included the alarmins TSLP and IL-33 to enable deeper endotyping of patients with T2 disease.

## Methods

### Patient Cohort and Clinical Phenotyping

The study protocol and design, including inclusion and exclusion criteria, was previously described.(10) Briefly, a cross-sectional study was conducted on patients with asthma recruited through the Yale Center for Asthma and Airway Diseases (YCAAD) between March 2009 and June 2017. Subjects were >12 years old, current nonsmokers, and asthmatic as diagnosed by National Asthma Education and Prevention Program guidelines. Recruitment and consenting procedures were approved by the Institutional Review Board and written informed consent was obtained from all participants. For this study, a total of 200 subjects were recruited. Clinical phenotyping entailed an asthma-focused history, physical examination, administration of an asthma questionnaire, PFTs with evaluation of FeNO and bronchodilator response, and laboratory testing of blood samples. Clinical data used for analysis were those recorded closest to the time of sputum induction. Missing clinical data are summarized in Table E2.

### Measurement of sputum cytokines

Sputum induction with inhaled hypertonic saline and sample processing (mucus plug isolation method) were performed as previously reported.(10) Sputum levels of 10 pre-selected cytokines including alarmins (TSLP and IL-33), T2 cytokines (IL-4, IL-5, IL-9, IL-13, IL-21), and non-T2 cytokines (GCSF, IL-1β, IL-6) were assessed. Measurements were performed using a customized U-PLEX Biomarker Multiplex Assay kit from Meso Scale Diagnostics (MSD; Rockville, MD) according to the manufacturer’s protocol. First, each of the 10 biotinylated antibodies was coupled to a unique linker, which enables immobilization of each primary antibody to a designated measurable spot within the wells of a 96-well U-PLEX plate. Linker-coupled antibodies were then mixed and attached to the plate by incubation at 4°C overnight with shaking. The plate was washed three times with phosphate buffered saline containing 0.05% Triton-X (PBS-T). 25 μl of diluent plus 25 μl of calibrator standard or sputum supernatant was added to each well, and the plate was incubated at room temperature (RT) for 1 hour with shaking, followed by three washes with PBS-T. Next, 50 μl of detection antibody solution was added to each well and incubated at RT for 1 hour with shaking. Finally, 150 μl of 2x Read Buffer was added to each well and the plates were read on the MESO 1300 QuickPlex SQ 120 (MSD; Rockville, MD). Sputum cytokine concentrations were determined by interpolation from their standard curve.

### Statistical Analysis

Undetectable concentrations were assigned a value of 0.01. Cytokine concentrations were then log-transformed and standardized. Table E1 summarizes the mean detectable cytokine concentration and the percentage of undetectable values for each cytokine. Associations between cytokines were examined using both Pearson’s correlation and hierarchical clustering. Highly correlated cytokines were grouped and referred to as ‘modules’. Associations between cytokines and clinical features were assessed using Spearman’s rank correlation due to the likelihood of non-linear relationships. Patients defined as having ‘Hi’ or ‘Lo’ module expression were those in the highest or lowest quartile of module expression, respectively. A Gaussian Mixture Model (GMM) implemented through the R package *mclust* was used to identify patient ‘clusters’ from sputum cytokine data.(18) The optimal cluster number was determined to be three based on optimality criterion generated by the *NbClust* package and through *post hoc* inspection of clustering plots. An ellipsoidal, equal volume, shape, and orientation (EEE) GMM was found to be optimal based on Bayesian Information Criterion. Throughout, continuous variables were compared using the Kruskal-Wallis test due to non-normal data distributions and presented as medians with interquartile range (unless otherwise indicated). Categorical variables were compared using the Chi-squared test and presented as frequencies and percentages. P-values of < 0.05 were considered statistically significant. Statistical analysis was conducted using the R software environment (V.3.6.0, Vienna, Austria).

## Results

A total of 200 patients were included in the analysis. Patient demographics, clinical features, PFTs, and sputum characteristics are presented in Table 1. The population was racially diverse and demonstrated a broad distribution of asthma severity. Consistent with prior studies of the YCAAD cohort, asthma patients had an elevated mean BMI and female predominance.

**Table 1:** Demographic, clinical, and sputum characteristics of patient clusters.

|  | Cluster 1 (N=46) | Cluster 2 (N=93) | Cluster 3 (N=61) | Total (N=200) | P-value |
| --- | --- | --- | --- | --- | --- |
| **Demographics and clinical variables, Median (IQR) unless specified** |  |  |  |  |  |
| **Age at Visit** | 51.0 (41.0, 61.0) | 44.0 (27.0, 54.0) | 49.5 (38.0, 61.5) | 48.0 (35.0, 57.5) | 0.019 |
| **Sex, n (%)** |  |  |  |  | 0.975 |
| Female | 31 (67.4%) | 61 (65.6%) | 40 (65.6%) | 132 (66.0%) |  |
| Male | 15 (32.6%) | 32 (34.4%) | 21 (34.4%) | 68 (34.0%) |  |
| **Race, n (%)** |  |  |  |  | 0.409 |
| American Indian or Alaska Native | 0 (0.0%) | 2 (2.3%) | 0 (0.0%) | 2 (1.0%) |  |
| Asian | 0 (0.0%) | 4 (4.5%) | 1 (1.6%) | 5 (2.6%) |  |
| Black | 10 (21.7%) | 18 (20.2%) | 13 (21.3%) | 41 (20.9%) |  |
| Other | 3 (6.5%) | 9 (10.1%) | 2 (3.3%) | 14 (7.1%) |  |
| White | 33 (71.7%) | 56 (62.9%) | 45 (73.8%) | 134 (68.4%) |  |
| **Hispanic Origin, n (%)** | 5 (11.4%) | 20 (23.5%) | 11 (18.3%) | 36 (19.1%) | 0.245 |
| **BMI** | 29.97 (24.59, 35.17) | 27.08 (23.57, 33.97) | 30.94 (26.61, 35.92) | 29.50 (24.34, 34.48) | 0.041 |
| **Age at Symptom Onset** | 24.0 (5.0, 45.8) | 11.0 (3.0, 28.0) | 22.0 (5.0, 36.0) | 18.0 (4.0, 37.0) | 0.030 |
| **Atopy, n (%)** | 34 (75.6%) | 83 (90.2%) | 51 (83.6%) | 168 (84.9%) | 0.076 |
| **Hospitalizations Within the Last Year, Mean (SD)** | 1.28 (2.41) | 0.28 (0.76) | 0.61 (1.29) | 0.61 (1.50) | 0.033 |
| **ACT Score** | 17.0 (11.8, 21.3) | 17.0 (11.0, 21.0) | 14.0 (10.8, 19.3) | 16.0 (11.0, 21.0) | 0.105 |
| **Serum IgE** | 198.0 (47.9, 396.0) | 216.0 (68.3, 374.5) | 130.0 (27.8, 300.8) | 149.0 (36.7, 359.5) | 0.364 |
| **Blood Eosinophils (x10<sup>3</sup>/μl)** | 321.0 (178.5, 456.8) | 231.0 (124.4, 437.5) | 195.0 (118.0, 360.0) | 250.0 (131.5, 435.5) | 0.164 |
| **Asthma Severity, n (%)** |  |  |  |  | 0.003 |
| Mild | 1 (2.27%) | 19 (22.62%) | 4 (6.67%) | 24 (12.77%) |  |
| Moderate | 15 (34.09%) | 29 (34.52%) | 27 (45.00%) | 71 (37.77%) |  |
| Severe | 28 (63.64%) | 36 (42.86%) | 29 (48.33%) | 93 (49.47%) |  |
| **Pulmonary Functions, Median (IQR)** |  |  |  |  |  |
| **Pre: FEV1 (% pred)** | 72 (55, 84) | 81 (60, 95) | 76 (63, 88) | 76 (60, 92) | 0.113 |
| **Pre: FVC (% pred)** | 83 (72, 96) | 92 (77, 103) | 86 (74, 98) | 89 (75, 101) | 0.130 |
| **Pre: FEV1/FVC (ratio)** | 0.66 (0.54, 0.78) | 0.72 (0.61, 0.80) | 0.73 (0.61, 0.79) | 0.71 (0.60, 0.79) | 0.191 |
| **Post: FEV (% pred)** | 77 (65, 91) | 87 (70, 98) | 83 (71, 96) | 83 (70, 97) | 0.126 |
| **Post: FVC (% pred)** | 89 (79, 102) | 94 (82, 105) | 91 (77, 103) | 92 (79, 104) | 0.217 |
| **Post: FEV1/FVC (ratio)** | 0.68 (0.60, 0.80) | 0.76 (0.65, 0.81) | 0.75 (0.68, 0.81) | 0.74 (0.64, 0.81) | 0.151 |
| **FeNO (ppb)** | 30.0 (13.3, 55.3) | 31.0 (18.3, 47.8) | 33.0 (20.0, 54.0) | 31.0 (18.0, 53.8) | 0.665 |
| **BDR (%)** | 7.61 (4.55, 21.91) | 6.67 (2.03, 12.51) | 8.57 (2.70, 14.83) | 7.61 (2.43, 14.43) | 0.425 |
| **Sputum Characteristics, Median (IQR); cell counts are given as % of total.** |  |  |  |  |  |
| **Total cell count (x10<sup>3</sup>/μl)** | 617.5 (261.3, 1030.0) | 640.0 (335.0, 1170.0) | 520.0 (240.0, 1050.0) | 630.0 (300.0, 1121.3) | 0.369 |
| **Neutrophils (%)** | 28.2 (18.0, 40.5) | 34.9 (15.7, 47.0) | 30.0 (23.0, 45.0) | 32.0 (17.9, 45.0) | 0.844 |
| **Eosinophils (%)** | 5.0 (2.2, 12.2) | 3.0 (1.8, 10.5) | 5.0 (2.0, 11.5) | 4.0 (2.0, 11.0) | 0.130 |
| **Macrophages (%)** | 58.9 (45.0, 69.0) | 54.0 (34.0, 71.8) | 58.0 (39.2, 69.2) | 56.1 (39.2, 70.1) | 0.592 |
| **Lymphocytes (%)** | 2.0 (1.0, 4.4) | 2.0 (1.0, 3.7) | 1.4 (1.0, 3.2) | 2.0 (1.0, 3.8) | 0.274 |

### Identification of cytokine signaling modules

Sputum cytokine levels were quantified using an ultrasensitive multiplexed electrochemiluminescent immunoassay as described in the Methods section. Correlations between cytokines were assessed by Pearson’s *r* and visualized in Figure 1. Three groups of highly correlated cytokines were identified: the TSLP module, or TSLP_mod_ (TSLP, IL-4, IL-5, IL-9); the IL-1β_mod_ (IL-1β, IL-6, GCSF); and the IL-33_mod_ (IL-33, IL-13, IL-21). The TSLP and IL-33 modules were named according to their characteristic alarmins. IL-1β was used to define its module because it similarly represents an upstream inflammatory mediator released in response to cellular stress.(22)

**Figure 1:**
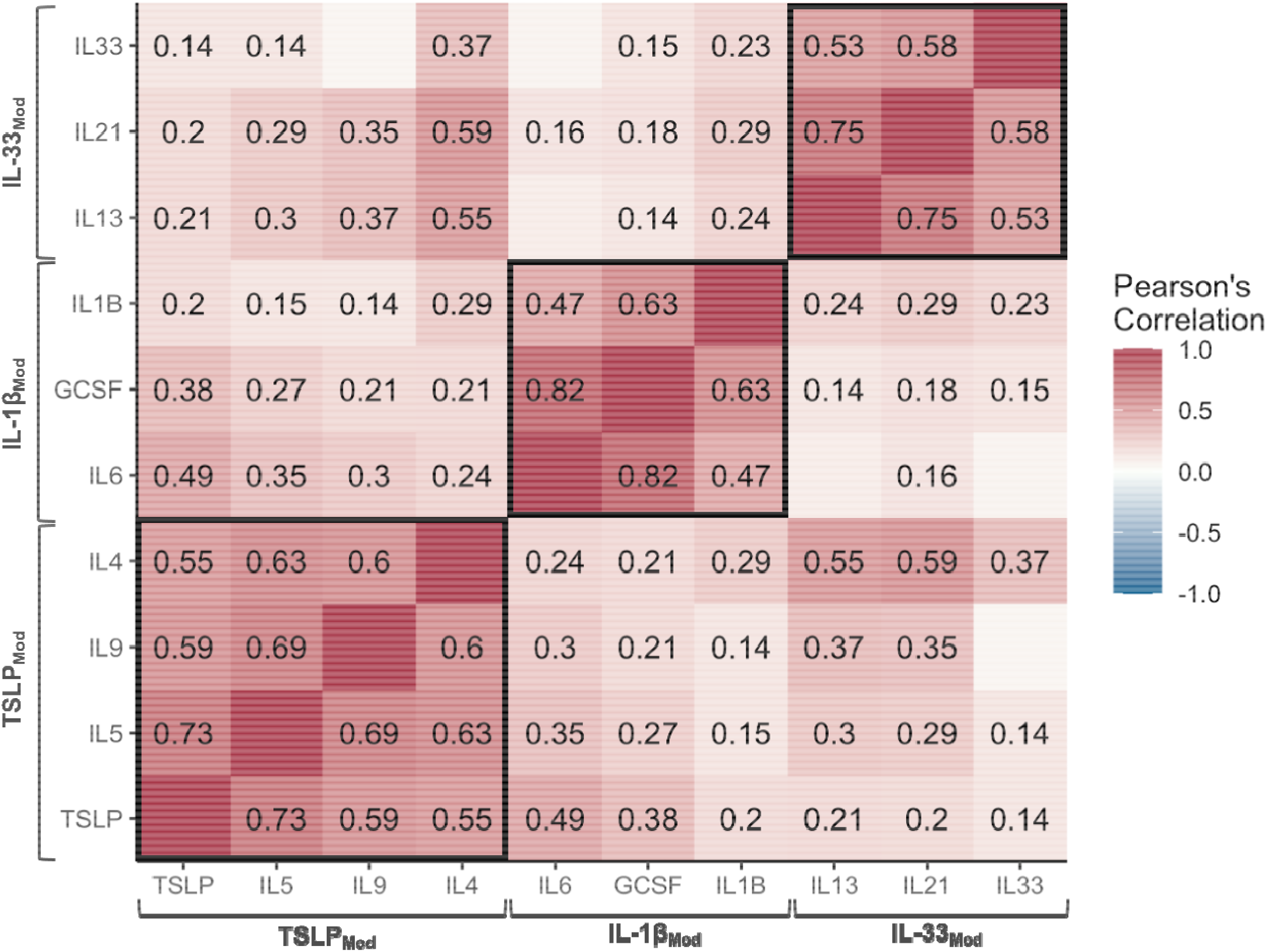
Correlations among sputum cytokines reveal three distinct signaling modules. Pearson’s correlation of detected cytokines; only significant correlation coefficients are shown. Three modules of correlated cytokines were identified: the TSLP_mod_ (TSLP, IL-4, IL-5, IL-9); IL-1β_mod_ (IL-1β, IL-6, GCSF); and IL-33_mod_ (IL-33, IL-13, IL-21).

The IL-1β_mod_ was consistent with a recognized non-T2 signaling pathway that includes IL-6 and GM-CSF, as described by Osei *et al*.(23) However, the canonical T2 cytokines (IL-4, IL-5, IL-13) did not correlate with each other as expected. Instead, two distinct T2 pathways were identified: the TSLP_mod_ (IL-4, IL-5, IL-9) and IL-33_mod_ (IL-13, IL-21). Notably, the alarmins TSLP and IL-33 did not correlate with each other, suggesting they may be expressed independently in the airways of patients with asthma.

To test the validity of the correlation-defined modules, we performed hierarchical clustering of cytokine levels. As shown in Figure E1, this unsupervised approach confirmed the associations between cytokines in each module. Of note, the three IL-33_mod_ cytokines all had relatively low levels (Table E1). To ensure that their association was not confounded by their frequency of undetectable values, we repeated the correlation analysis using only measurable values (Fig. E2). This analysis demonstrated similar correlations between IL-33, IL-13, and IL-21, confirming their characterization as a distinct cytokine signaling module.

In order to identify clinical features associated with each module, we compared patients with high expression to those with low expression of each module (Table E3). The most striking differences were observed in TSLP_mod_^Hi^ patients, who had more severe disease (p < 0.05), frequent hospitalizations (p = 0.06), lower lung function (p < 0.01), higher sputum and blood eosinophil counts (p < 0.05), and higher FeNO levels (p < 0.001). Notably, this population had significantly lower sputum neutrophil levels (p < 0.05). In contrast, IL-1β _mod_^Hi^ patients showed a trend towards higher sputum neutrophils. The only significant finding in IL-33_mod_^Hi^ patients was a lower ACT score, indicating poorer disease control (p < 0.05).

To identify associations between clinical parameters and individual cytokines, we assessed Spearman’s correlation. As shown in Figure E3, TSLP correlated significantly with several markers of clinical severity, indicating that it may be the central driver of the TSLP_mod_^Hi^ phenotype. Indeed, a recent trial demonstrated that monotherapy with the TSLP inhibitor Tezepelumab was sufficient to produce improvements in all of the same clinical parameters.(24)

### Identification of patient subgroups

Next, patients were clustered according to sputum cytokine expression using LCA. The optimal solution was achieved using three clusters with a EEE GMM.(25) To visualize the results, contour plots of estimated mixture density of the clusters were projected on a dimension-reduced subspace (Fig. 2).(26) This analysis demonstrated two distinct, but related clusters (1 and 2), and clear separation of a third cluster (3).

**Figure 2:**
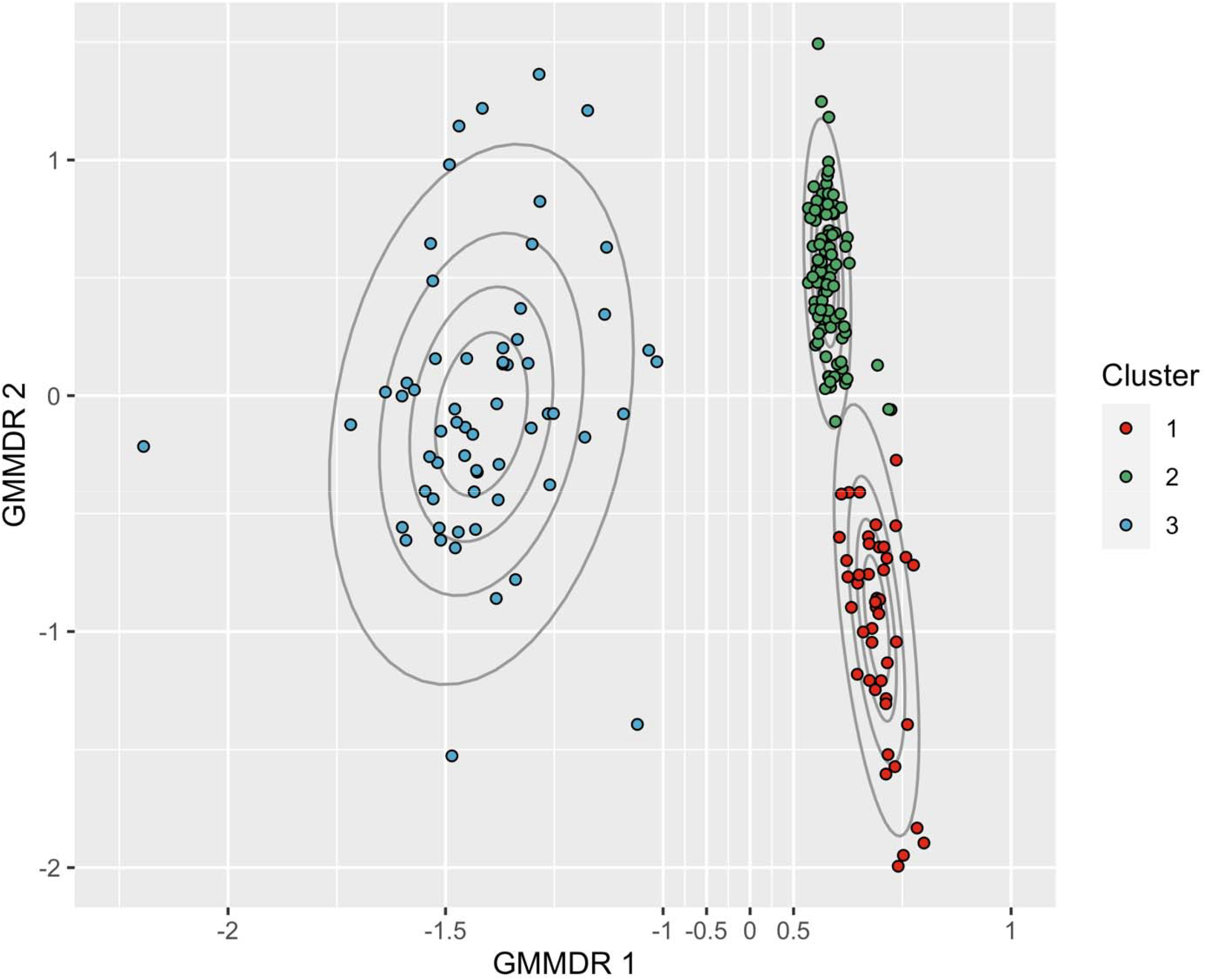
Clustering of patients by sputum cytokine levels reveals three cytokine subgroups. Latent class analysis was used to cluster patients according to airway cytokine expression. Contour plots of the three resulting clusters are represented.

Cytokine profiles of each patient are presented in the heatmap in Figure 3, and the expression of cytokine modules among the patient clusters is shown in Figure 4. Cluster 1 showed an isolated increase in the TSLP_mod_; cluster 2 showed decreased expression of both T2 modules; and cluster 3 had the highest expression of all three modules. Interestingly, cluster 1 showed low IL-33_mod_ together with high TSLP_mod_ expression, suggesting differential regulation of these modules in a subgroup of patients with asthma. In summary, this analysis suggests that alarmins, T2, and non-T2 cytokines are differentially expressed among patients with asthma and may contribute to heterogeneity of disease.

**Figure 3:**
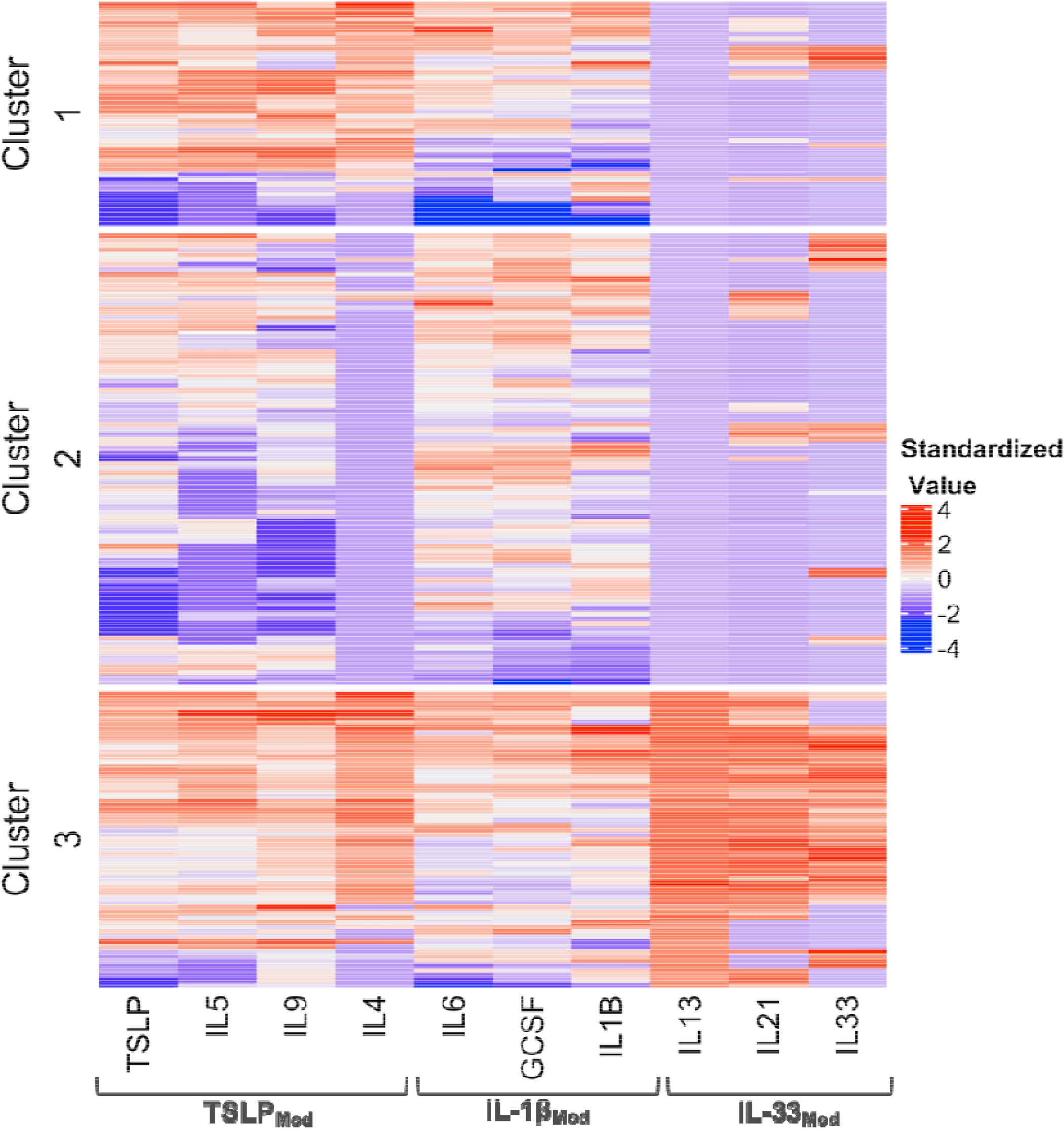
Heatmap of cytokine profiles of patients within each cytokine cluster. A hierarchical clustering heatmap of sputum cytokine levels is presented. Cytokine profiles are organized by cluster, demonstrating the variability of cytokine levels across the study population.

**Figure 4:**
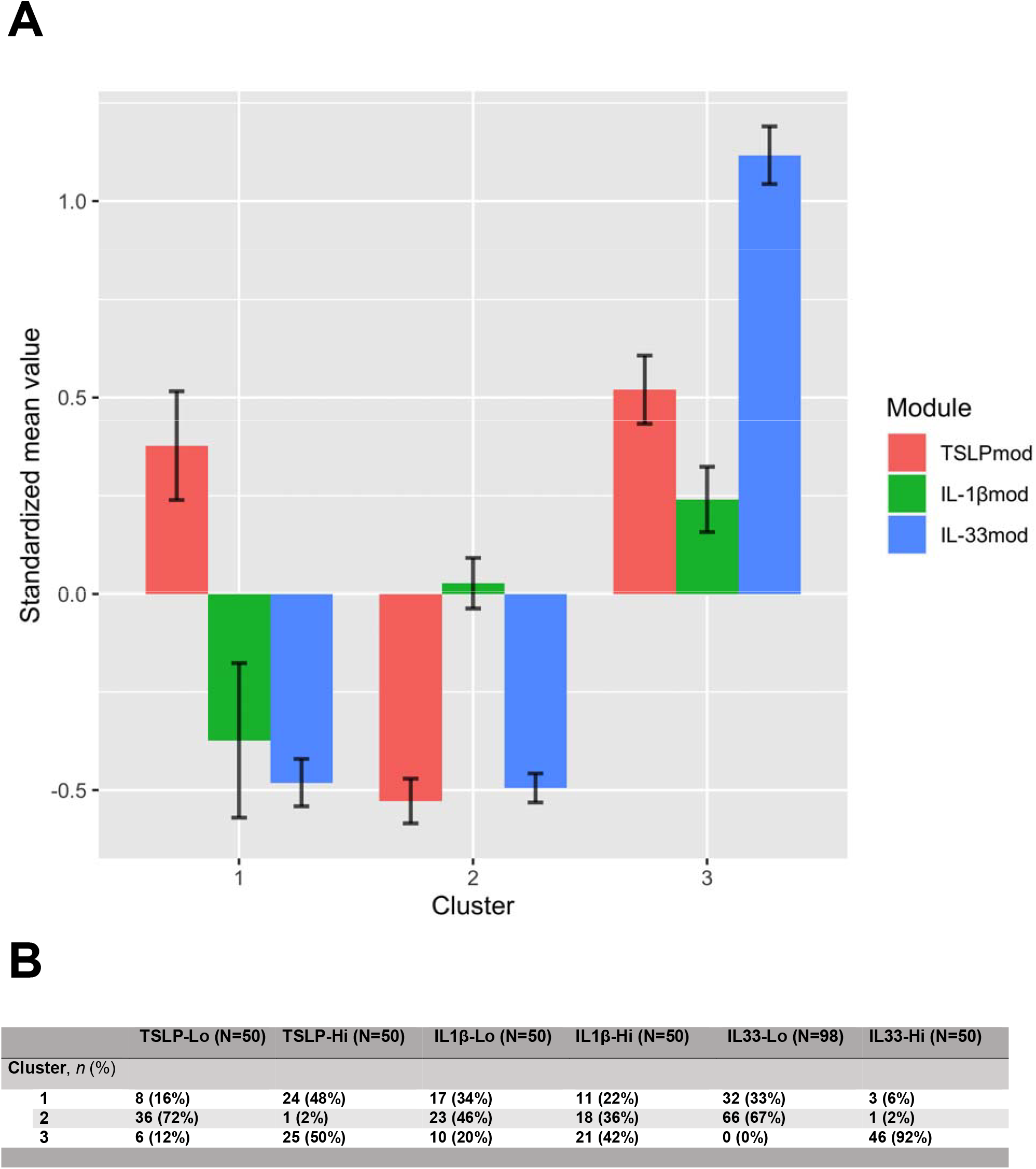
Expression of signaling modules within each cytokine cluster. **A**, Latent cytokine profiles based on standardized mean value were grouped by module and averaged. Cluster 1 shows isolated expression of TSLP_mod_. Cluster 2 shows depression of the TSLP_mod_ and IL-33_mod_. Cluster 3 shows global increases in module expression. **B**, The distribution of patients with high module expression as defined in Table E3 are presented within the context of the clusters.

Finally, we compared the demographics, clinical traits, PFTs, and sputum characteristics between the clusters (Table 1). Consistent with the phenotype of TSLP_mod_^Hi^ patients in Table E3, cluster 1 demonstrated severe disease and frequent hospitalizations. Patients in cluster 2, who showed the lowest expression of pathogenic cytokines (Fig. 4), had younger age of onset, lower BMI, less frequent hospitalizations and milder disease severity. Cluster 3 showed no distinguishing clinical features.

## Discussion

The T2 vs non-T2 paradigm represents an attractive framework for defining asthma pathophysiology and personalizing therapy (Fig. 5A). However, it has proven inadequate to predict patient response to targeted therapy. Indeed, up to 50% of patients with classic T2 features such as eosinophilia fail to respond to biologic agents against T2 cytokines.(8) Thus, there appear to be additional subtypes of disease that cannot be identified using existing diagnostics.

**Figure 5:**
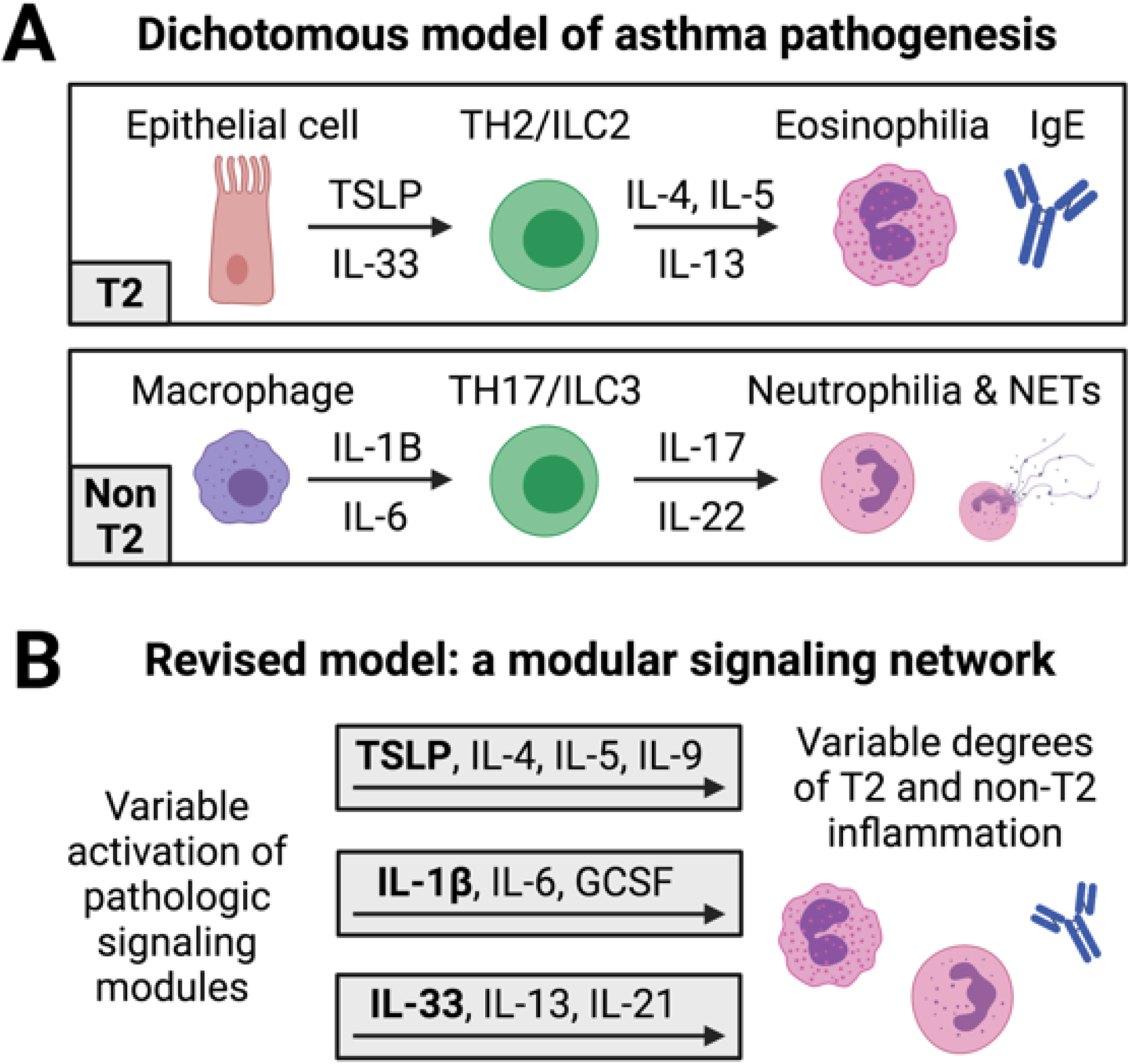
Schematic of pathologic signaling pathways in asthma. **A**, A dichotomous model of asthma pathogenesis, driven by either T2 or non-T2 immune responses. **B**, A revised model with three independent signaling modules, each variably activated in patients with asthma to produce unique biological endotypes. These modules contribute to varying degrees to produce a clinical phenotype that may have both T2 and non-T2 features. Figure created with BioRender.com.

To address this issue, we evaluated whether patients could be delineated more effectively using airway alarmin and cytokine levels. To this end, we quantified sputum cytokine expression in patients with asthma, defined signaling modules, and assessed expression of these modules in unique patient subgroups. In this analysis, we included the alarmins TSLP and IL-33, important pathogenic mediators in asthma that have not been included in clustering studies to date.(13) In addition, to address the problem of low-abundance cytokines such as IL-33 (Table E1), we utilized an electrochemiluminescence-based immunoassay, which offers 10-fold higher sensitivity than standard assays.

Using this novel approach, we identified two distinct T2 signaling pathways, each associated with a unique alarmin: the TSLP_mod_ and IL-33_mod_. As demonstrated by the patients in cluster 1, who showed increased TSLP_mod_ and depressed IL-33_mod_ expression, these pathways may be regulated independently. This finding represents some of the first human evidence for mechanistic heterogeneity within the T2 asthma paradigm, along with the transcriptomic work of Peters et al.(11) The existence of T2 subtypes has been inferred from the ability to substratify T2 patients by atopic status, and from the variable clinical response to T2 biologics,(17, 27, 28) but the molecular basis of these subtypes has remained largely unknown. Further translational ‘omics and pre-clinical studies should offer additional insights into this incipient area of research.(29, 30)

The identification of these immune correlates of T2 heterogeneity could have important therapeutic implications. For instance, TSLP_mod_^Hi^ patients may respond well to anti-IL-5 biologics, while IL-33_mod_^Hi^ individuals may respond better to medications targeting IL-13 or IL-33, such as astegolimab.(31) Furthermore, synergism may be achieved using combination therapy against multiple cytokines within a given module, such as TSLP and IL-5. Prospective trials will be needed to test these hypotheses, but the targeting of pathogenic cytokines based on sputum levels could represent a rational, mechanistic-based strategy to improve rates of clinical response.

Our results also provide evidence for a non-T2 signaling mechanism defined by IL-1β, IL-6, and GCSF (the IL-1β_mod_). This pathway is consistent with transcriptomic studies implicating inflammasome activation in non-T2 asthma,(17, 32) and sputum analyses showing elevated IL-1β levels together with caspase activity, neutrophilic inflammation, and neutrophil extracellular traps.(33, 34) However, IL-1β_mod_ signaling was not associated with a distinct non-T2 endotype in our study. Instead, IL-1β_mod_ cytokines were co-expressed with T2 cytokines in cluster 3. This observation argues against a dichotomous T2 vs non-T2 model, suggesting that patient pathobiology may be better described as a network of multiple independent T2 and non-T2 signaling modules (Fig. 5B). It is also interesting to note that the global upregulation of cytokines in cluster 3 was not associated with increased asthma severity, indicating that patient pathology may derive from imbalanced module expression rather than absolute increases in cytokine production.

Importantly, the cytokine-based clusters described here did not correlate with readily apparent clinical phenotypes or currently used biomarkers. Cluster 3 patients, for example, possessed a highly distinct immunologic signature compared to clusters 1 and 2 (Fig. 3), but no defining clinical features. This observation underscores the inability of current clinical metrics to delineate immunologic endotypes of asthma, and suggests that sputum proteomics could represent an effective alternative.

Limitations of this study include the relatively narrow set of cytokines analyzed, which was constrained by experimental feasibility and the number of cytokines targetable by existing biologics. Unbiased proteomic analysis, in contrast, would have enabled deeper characterization of airway immune responses and greater opportunity for pathophysiologic discovery. In addition, despite the substantial size and diversity of the study population, our analysis lacks a validation cohort. As such, follow-up studies will be necessary to confirm the generalizability of our findings. Finally, the cross-sectional design of this study precludes conclusions regarding the stability of disease subtypes over time, and regarding the mechanistic roles of the signaling modules identified. Further evaluation in translational studies and preclinical models will be needed to evaluate these questions.

Acknowledging these caveats, we believe that the alarmin-based modular signaling framework proposed herein merits consideration as a model of asthma pathophysiology. We also believe that a therapeutic strategy based on sputum cytokine profiling could represent a practical, cost-effective, and rational approach for selecting biologics for patients with asthma – a significant step towards the implementation of precision medicine for this common yet highly complex disease.

## Data Availability

All data produced in the present study are available upon reasonable request to the authors.

## Abbreviations

ACT: Asthma control test
BDR: Bronchodilator response
BMI –: Body mass index
FeNO: Fractional exhaled nitric oxide
IgE: Immunoglobulin E
LCA: Latent class analysis
PFT: Pulmonary function test
T2: Type 2
TSLP: Thymic stromal lymphopoietin
YCAAD: Yale Center for Asthma and Airway Diseases

## Supplemental figure legends

**Figure E1:**
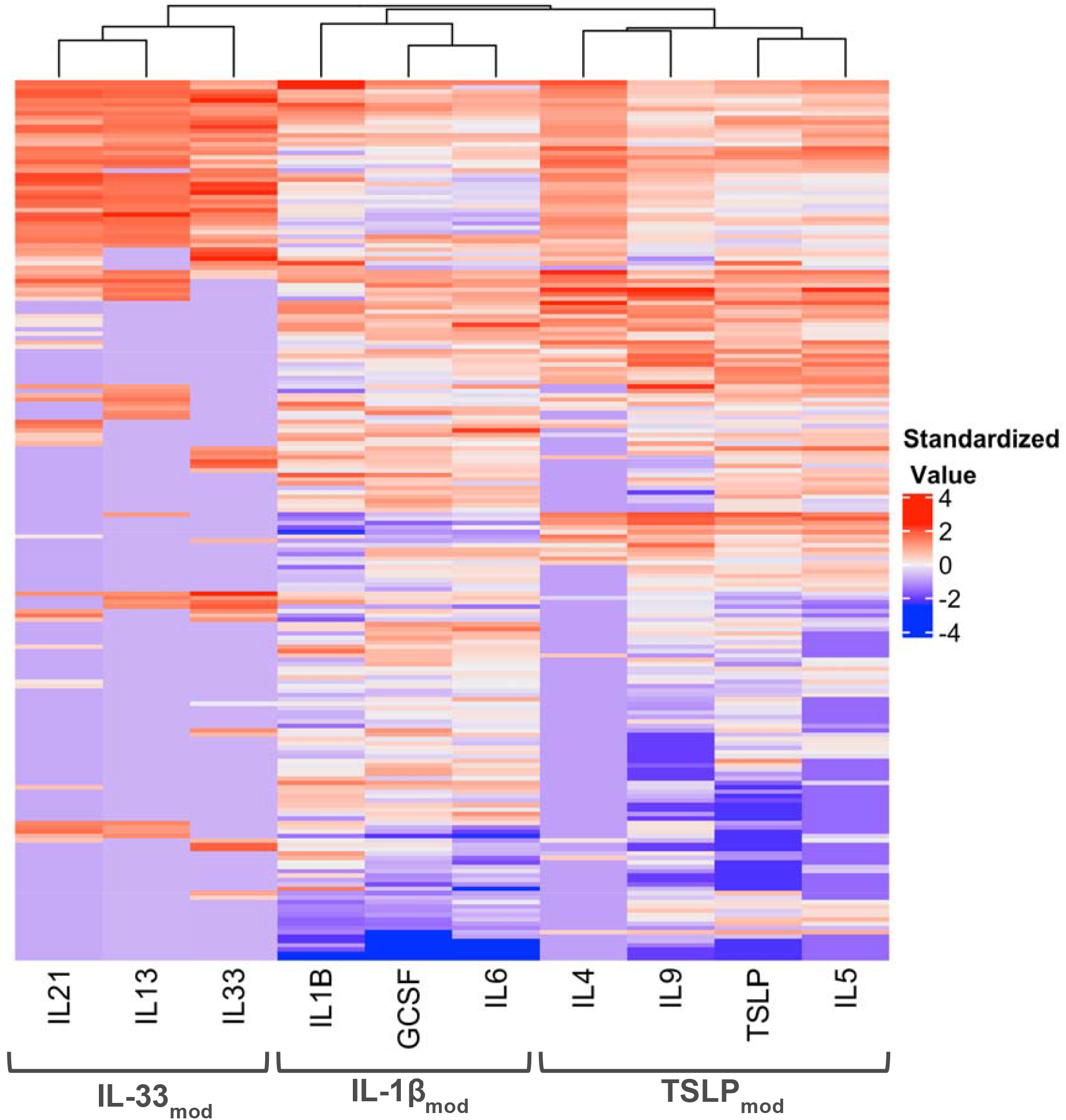
Hierarchical clustering of sputum cytokines confirm three distinct signaling modules. Clustering analysis confirms the cytokine modules determined by Pearson’s correlation in Figure 1.

**Figure E2:**
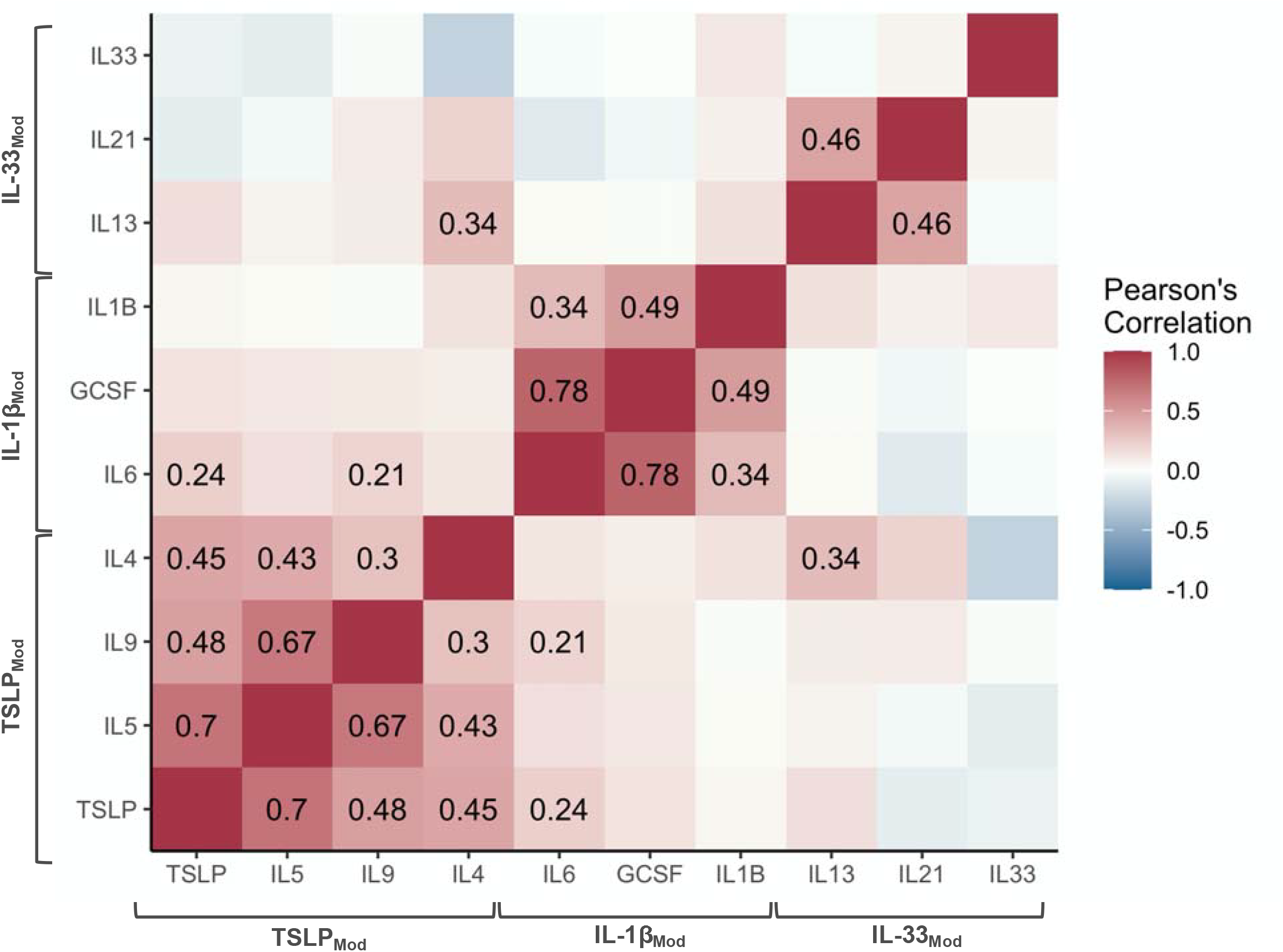
Correlations among sputum cytokines with detectable values confirm the existence of three distinct signaling modules. Pearson’s correlation was performed specifically on measurable cytokine values to confirm the validity of the correlations shown in Figure 1.

**Figure E3:**
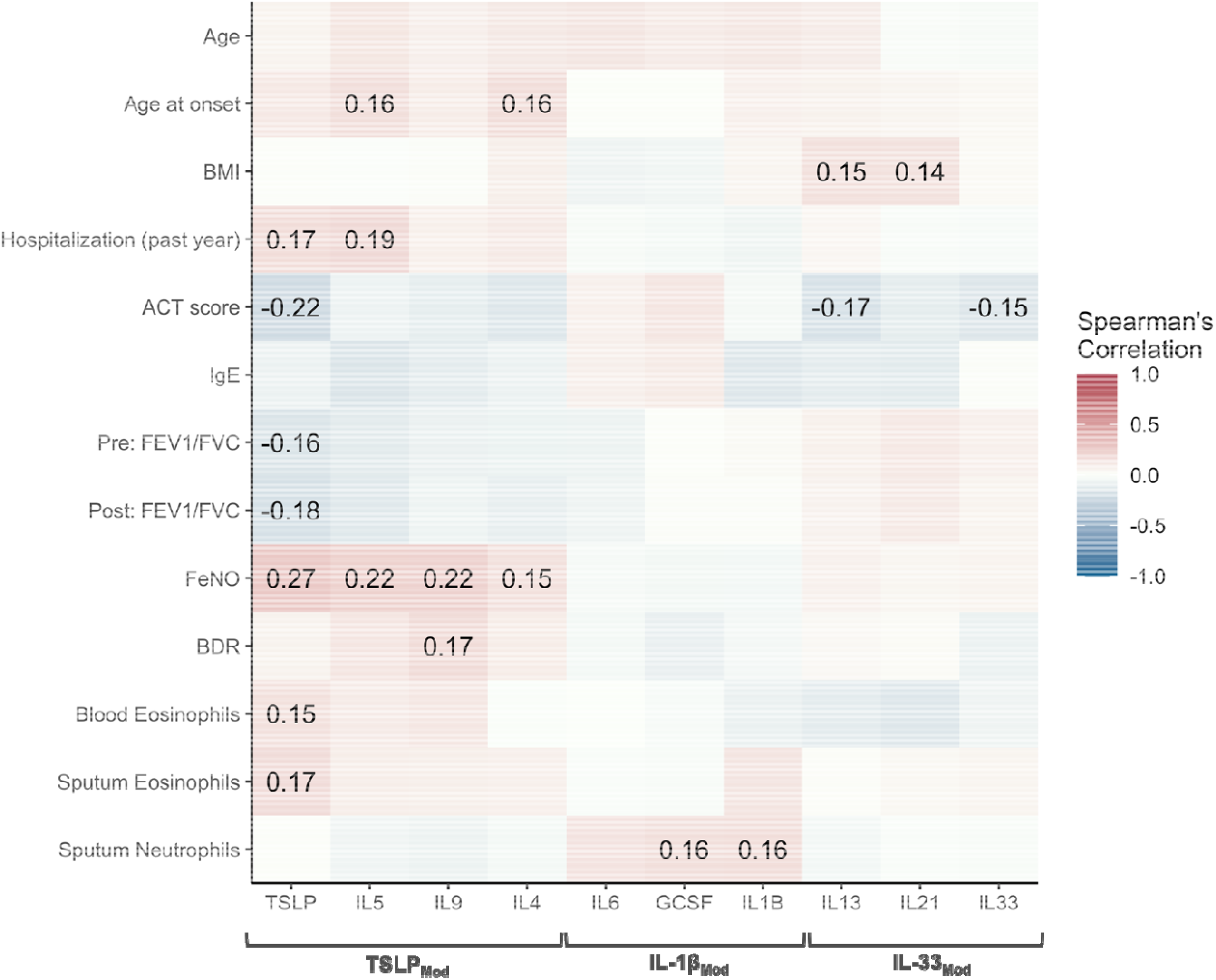
Correlations between cytokines and selected clinical features. Spearman’s correlation was performed on individual cytokines and clinical parameters.

**Table E1:** Standardized mean values and percent undetectable of sputum cytokines.

| | TSLP | IL-5 | IL-9 | IL-4 | IL-6 | GCSF | IL-1β | IL-13 | IL-21 | IL-33 |
| --- | --- | --- | --- | --- | --- | --- | --- | --- | --- | --- |
| **Mean Detectable Value (pg/mL)** | 2.84 | 10.83 | 6.97 | 0.74 | 82.12 | 107.57 | 102.79 | 22.84 | 1.35 | 3.55 |
| **Undetectable, *n* (%)** | 23 (12%) | 48 (24%) | 21 (10%) | 106 (53%) | 6 (3%) | 7 (4%) | 2 (1%) | 139 (70%) | 121 (60%) | 132 (66%) |

**Table E2:** Missing clinical data characterization.

| **Parameter** | **Missing Data, *n* (%)** |
| --- | --- |
| Age at Visit | 1 (0.5%) |
| Race | 4 (2%) |
| Hispanic Origin | 11 (5.5%) |
| BMI | 1 (0.5%) |
| Age at Symptom Onset | 1 (0.5%) |
| Atopy | 2 (1%) |
| ACT Score | 11 (5.5%) |
| Serum IgE | 96 (48%) |
| Blood Eosinophils | 17 (8.5%) |
| Asthma Severity | 12 (6%) |
| Pre: FEV1 (% pred) | 3 (1.5%) |
| Pre: FVC (% pred) | 3 (1.5%) |
| Pre: FEV1/FVC (ratio) | 3 (1.5%) |
| Post: FEV (% pred) | 3 (1.5%) |
| Post: FVC (% pred) | 3 (1.5%) |
| Post: FEV1/FVC (ratio) | 3 (1.5%) |
| FeNO | 14 (7%) |
| BDR | 3 (1.5%) |
| Total cell count | 4 (2%) |
| Viability | 2 (1%) |
| Neutrophils | 23 (11.5%) |
| Eosinophils | 35 (17.5%) |
| Macrophages | 14 (7%) |
| Lymphocytes | 50 (25%) |

**Table E3:**
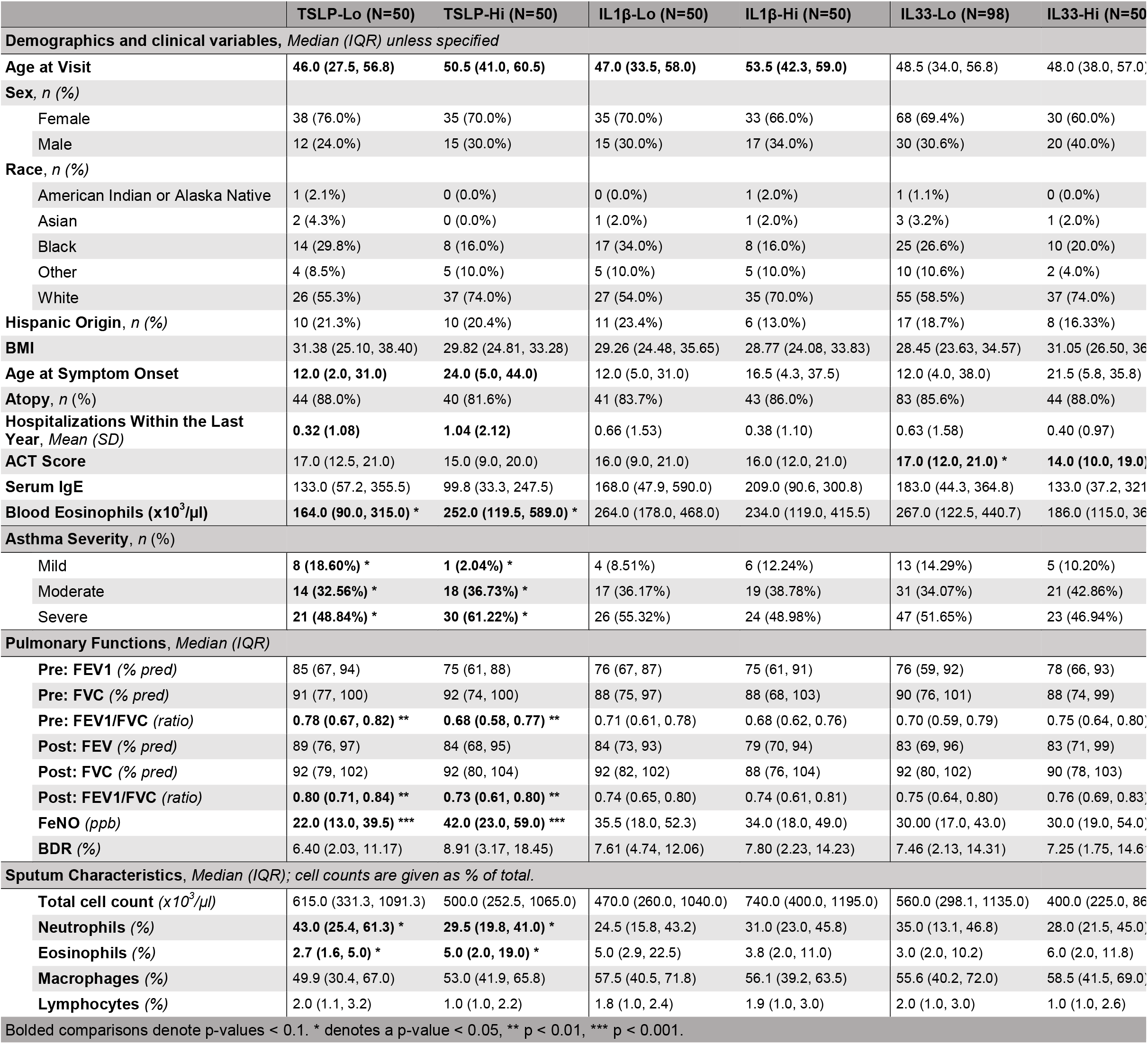
Demographic, clinical, and sputum characteristics of patients grouped according to module expression.

